# Linking brain maturation and puberty during early adolescence using longitudinal brain age prediction in the ABCD cohort

**DOI:** 10.1101/2022.05.16.22275146

**Authors:** Madelene C. Holm, Esten H. Leonardsen, Dani Beck, Andreas Dahl, Rikka Kjelkenes, Ann-Marie G. de Lange, Lars T. Westlye

## Abstract

The temporal characteristics of brain maturation could potentially represent a mediating effect between pubertal development and life outcomes. Using a large longitudinal dataset of children aged 9-12 from the Adolescent Brain Cognitive Development (ABCD) study we tested the associations between pubertal status and brain maturation. Brain maturation was assessed using brain age prediction with a deep learning approach based on convolutional neural networks and minimally processed T1-weighted structural MRI data. Brain age prediction provided highly accurate and reliable estimates of individual age, with an overall mean absolute error of 0.7 and 1.4 years at the two timepoints respectively, and an intraclass correlation of 0.65. Linear mixed effects (LME) models accounting for age and sex showed that on average, advancing pubertal development by one pubertal stage was associated with a 2.4 months higher brain age across time points (β= 0.10, p<.001). Further, significant interactions with time demonstrated that higher rates of pubertal development were associated with larger positive changes in brain age over time (p<.001). These results demonstrate a link between sexual development and brain maturation in early adolescence, and provides a basis for further investigations of the complex sociobiological impacts of puberty on the adolescent brain and mind.

## Introduction

Brain development during adolescence is characterized by a highly coordinated sequence of both progressive (cell growth and myelination) and regressive (synaptic pruning) processes (Paus et al., 2008), observable as nonlinear trajectories of cortical thinning and white matter volume increase in relation to chronological age (Blakemore & Choudhury, 2006). The neurodevelopmental progress is most likely shaped by a complex interplay of genetic factors, changes in biological processes, and new environmental pressures (Fernandez-Cabello et al., 2022; Ferschmann et al., 2022). In parallel to brain development, adolescence is a period of drastic changes in physiological processes and body composition during puberty. Puberty refers to the reactivation of the hypothalamic pituitary gonadal axis that has remained dormant since early postnatal life, causing a steep increase in circulating gonadal steroids such as estradiol, progesterone, and testosterone (Campbell et al., 2009). The heightened levels of gonadal steroids primarily drive maturation of reproductive systems and secondary sex characteristics but puberty has also been linked to cortical (Vijayakumar et al., 2021) and subcortical gray matter (GM) (Goddings et al., 2014; Wierenga et al., 2018), as well as white matter (WM) (Blakemore et al., 2010) brain maturation. Moreover, rodent studies have showed neurotrophic and neuroplastic effects of estrogen and testosterone (Hsu et al., 2003; Filová et al., 2013), and human longitudinal studies have linked endogenous estrogen exposure to beneficial effects on brain ageing (Schelbaum et al., 2021), supporting a direct effect of gonadal hormones on brain structure throughout the human lifespan in women (Galea et al., 2027; de Lange et al., 2020). Given this evidence, puberty is a forefront candidate for biological processes shaping brain development during adolescence in addition to genetically programmed change.

The timing of puberty onset differs between the sexes with 1 year on average, such that females start their pubertal development between the ages 8 and 12, and males between ages 9 and 14 (Campbell et al., 2009). This one-year difference has been linked to the disproportion of depressive disorders, and depressive/internalizing symptoms, in young women (Pfeifer & Allen, 2021). Earlier onset of puberty has also been linked to positive effects such as higher academic achievements both in boys and girls, and may partly explain sex differences in educational achievement (Torvik et al., 2021). Although the mechanisms explaining sex differences in school performance are highly complex and multifaceted, it is conceivable that individual differences in brain maturation in early school years represent a relevant predictor for later life outcomes.

Related to their head start in puberty maturation, it is likely that females, at the group level, differ in the temporal characteristics of adolescent brain maturation compared to males. Early attempts to elucidate relevant sex-differences in specific brain structures during adolescence have yielded inconclusive and often contradicting findings (Lenroot & Giedd, 2010), which might be due to the narrow focus on predefined brain structures. In contrast, evidence points towards individually varying, mosaic compositions, of male/female like brain regions (Joel & Fausto-Sterling, 2016). Thus, recent studies have adopted a broader approach and moved towards multivariate integration of brain structures when distinguishing between male and female brain morphology (Brennan et al., 2021), multimodal investigations (Kaczkurkin et al., 2019), and investigating other characteristics of neurodevelopmental trajectories, such as sex differences in variability in brain structure (Wierenga et al., 2018a; Wierenga et al., 2019). As such, while the overall sex differences in brain morphology may be relatively small, the temporal characteristics of brain development during sensitive periods, such as puberty, may show additional relevant individual differences related to sexual development during adolescence.

In this study, in order to assess the overall relationship between pubertal development and brain maturation, and to investigate potential sex differences in brain maturational tempo, we linked pubertal development to early adolescents’ brain age based on brain structural MRI. Pubertal development was assessed using parent-reported development of physical secondary sex characteristics (the Pubertal Developmental Scale (PDS); Petersen, 1988), and brain maturation was assessed using brain age prediction based on brain MRI.

Brain age is a machine-learning based estimate of an individual’s age based on their brain structural features. Supervised algorithms are trained to learn age-related patterns in brain structure from a large set of brain images with a wide age span, and can subsequently be applied to unseen datasets to predict age at the individual level. In adults, the difference between brain age and chronological age has been shown a to represent a heritable trait (Kaufmann et al., 2019; Cole et al., 2017) and a range of clinically relevant characteristics and conditions have been associated with higher brain age (Tønnesen et al., 2020; Beck et al., 2022ab; de Lange et al., 2020; Høgestøl et al., 2019). In children and adolescents, brain age has been interpreted as an indicator of overall level of brain maturation (Franke et al., 2012; Brown et al., 2012). Longitudinal assessments in adolescence have shown brain age to be heritable, higher in females than males (Brouwer et al., 2021), and linked to psychopathology and psychosocial functioning (Drobinin et al., 2021; Cropley et al., 2021). However, the clinical and functional correlates of brain age in adolescence have not been fully established.

In the current study, we calculated brain age using a recent deep learning approach based on convolutional neural networks (CNNs) in a large training set comprised of minimally processed T1-weighted MRI data from >50,000 individuals aged 5 to 93 years (Leonardsen et al., 2022). We applied the model to predict brain age in the Adolescent Brain Cognitive Development (ABCD) cohort, including 7459 baseline scans and 2384 scans from the first MRI follow-up two years later.

Based on current models and studies reviewed above, we expected cross-sectional and longitudinal associations between pubertal development and estimated brain age over and above chronological age. Specifically, independent of age, we hypothesized that 1) participants rated with more advanced puberty development would show higher brain age across time points. Next, we hypothesized that 2) higher rate of longitudinal pubertal development between time points would be associated with higher rate of brain age change. Furthermore, based on a recent report (Brower et al., 2021) we expected 3) higher brain age in females compared to males, likely explained by sex differences in pubertal development.

## Methods

### Sample characteristics

We used data from the ongoing longitudinal ABCD study, where more than 11000 participants and their parents/guardians will be followed for ten years, with MRI data collection every second year (Garavan et al., 2018). Data used in the present study were downloaded in March 2022 as part of the ABCD Study Curated Annual Release 4.0 containing data from baseline up until the second-year visit (https://data-archive.nimh.nih.gov/abcd). To minimize confounding effects from complex family-related factors we included one participant per family for analysis. We excluded participants with known prenatal drug exposure, any serious medical, psychiatric, neurodevelopmental disorder and/or substance abuse, resulting in N=7459 (3987 female) at timepoint 1, and N=2384 (1239 female) at timepoint 2. The age distribution of the sample can be seen in figure 1.

**Figure 1.**
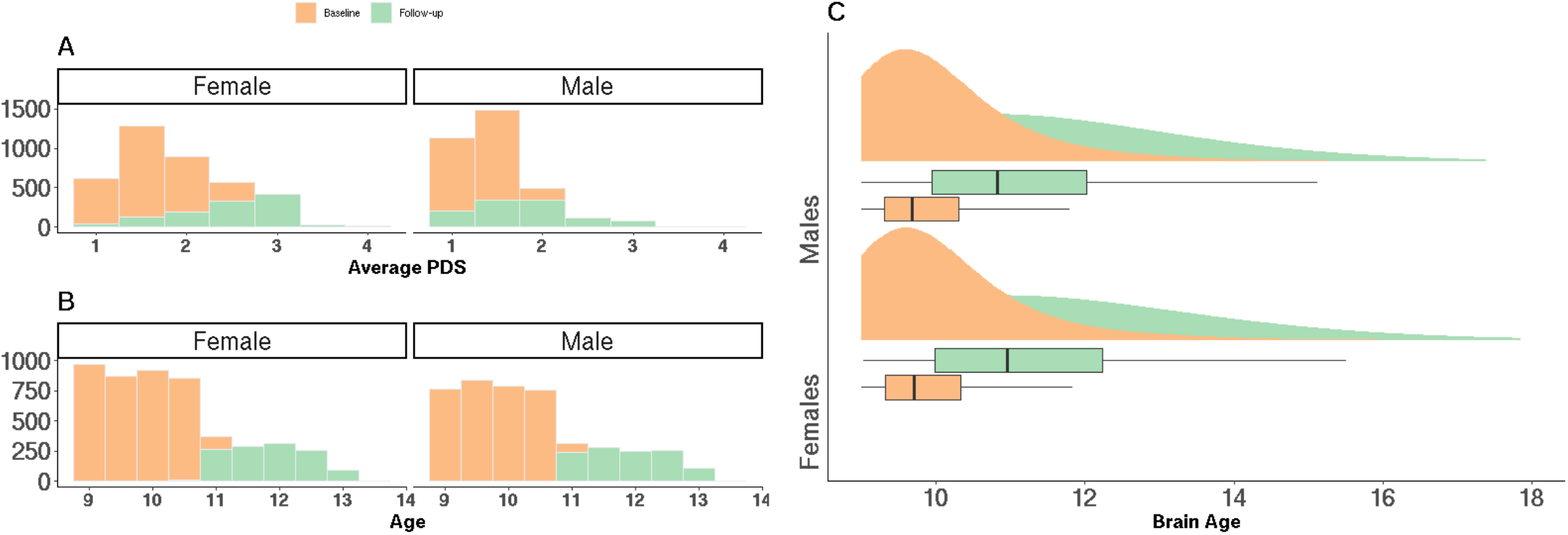
A: Sample distribution of average PDS at baseline and follow up. B: Age distribution at baseline and follow-up. C: Brain age distribution at baseline and follow up in females and males separately.

### Ethical approval

A centralized institutional review board approval of procedures was obtained from the University of California, San Diego. Written informed consent was obtained by parent or guardian, and assent from the participants, before partaking in the ABCD study. The current study has been approved by the Regional Committees for Medical and Health Research Ethics South-East Norway.

### MRI acquisition and processing

T1-weighted images were acquired with real time motion correction and imaging parameters harmonized for three 3T scanner platforms (Siemens Prisma, General Electric (GE) 750 and Philips) (Casey et al., 2018) and minimally processed (skull stripping, reorientation, and normalization) as described in detail in Leonardsen et al., (2022).

### Brain age calculations

The estimated brain age for each participant was calculated using a CNN trained and validated in minimally processed T1-weighted MRI data (n=53542, 5-93 years) from 21 publicly available datasets (Leonardsen et al., 2022). The model architecture is a regression variant (SFCN-reg) of the PAC2019-winning SFCN model (Peng et al., 2021). The model was trained, optimized and validated in subsets of the data (n=34285 and 8455 respectively) and achieved a validation mean absolute error (MAE) of 2.51. More importantly, an MAE of 2.47 was observed in a subset of the original data containing previously unseen participants, and an MAE of 3.90 in an external dataset from unknown scanners indicates exceptional generalization properties.

### Pubertal development assessment

Pubertal development was measured using PDS, a self or parent-rated questionnaire designed to mimic traditional Tanner staging assessment without the use of reference pictures (Petersen, 1988), in which puberty related development of physical secondary sex characteristics are ordinally rated. The questionnaire consists of seven items of which three are sex neutral, assessing skin changes, body hair changes, and growth in height. Two items are specific for females which are breast development and menarche (first menstruation), and two items are specific to males which are voice changes and facial hair growth. All items are rated on a scale of 1-4 (1: has not yet begun, 2: has barely begun, 3: is definitely underway, and 4: seems complete). The exception is the menarche item, which is a binary response item. Both parent and child ratings are available. PDS has shown high inter-rater reliability between both parent and self-rated assessment to clinicians, and correlates highly with plasma levels of gonadal hormones (Koopman-Verhoeff, 2020; Carskadon & Acebo, 1993). For analysis, we used parent-rated development scores, which have generally been shown to have higher correspondence with trained clinician assessments than child-rated scores (Rasmussen et al, 2015). Average scores across items were calculated for analysis purposes. The sample distribution of average pubertal development can be seen in figure 1.

### Statistical analysis

All statistical analysis were implemented in R version 4.0.0 (R core team, 2021). We employed LME models of varying complexity to test our hypotheses, using the lmerTest package v. 3.1-3 (Kuznetsova et al., 2017).

To assess our first hypothesis of an overall association between pubertal status and brain age, we tested two LMEs including estimated brain age as dependent variable and average PDS scores as predictive variables: one only controlling for age, and one controlling for sex and age. The models used average PDS, time point, age and sex as fixed effects, and scanner site and subject ID as random effects.

These models were defined as:

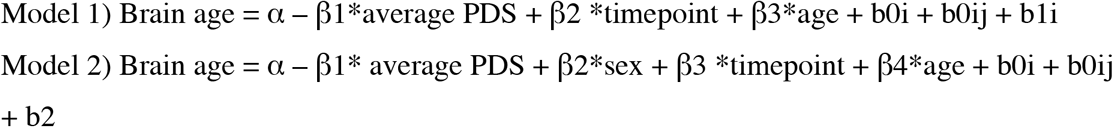

Where α denotes the intercept, β’s denoting the fixed effects slopes for sex, average PDS scores, age, and change over timepoints 1 and 2, and b0’s denoting random intercepts for subjects i and scanner j, and b1i denoting random slopes for average PDS per participants.

Next, to assess our second hypothesis that the rate of longitudinal changes in pubertal development is associated with longitudinal changes in brain age, we tested for interactions between average PDS and timepoint with predicted age as outcome. The model included sex, averaged PDS score, age, and timepoint as fixed factors, and subject ID and scanner site as random factors.

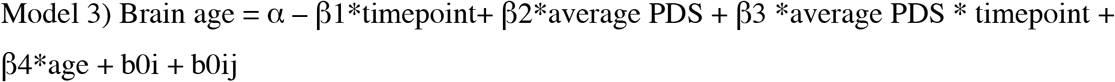

Where α denotes the intercept, β’s denoting the fixed effects slopes for sex, averaged PDS scores, age, and change over timepoints 1 and 2, and b’s denoting random intercepts for subjects i and scanner j.

Our third hypothesis of sex differences in brain age was tested in three models. One model assessed the main effect of sex, including only sex as predictor variable, controlling for age. The second model tested for sex differences in longitudinal change in brain age with an interaction term between sex and time. The third model tested for an interaction between sex and average PDS to assess whether the males and females differ in their pubertal effects on brain maturation.

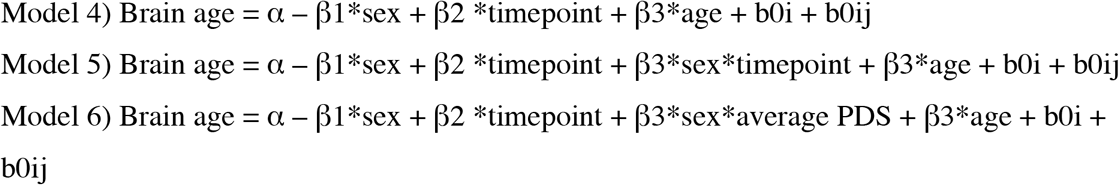

Where α denotes the intercept, β’s denoting the fixed effects slopes for sex, average PDS scores, age, and change over timepoints 1 and 2, and b’s denoting random intercepts for subjects i and scanner j, and random slopes for average PDS per participants.

All resulting p-values were corrected for multiple comparisons by false discovery rate using the p.adjust function in R (R core team, 2021).

## Results

### Descriptive statistics of pubertal development

Figure 1 shows the distribution of average PDS score in males and females. LMEs revealed significantly higher average PDS in females (mean=2.09, SD=0.65) compared to males (mean=1.52, SD=0.44, t=46.35, p<.001) across timepoints. The models revealed a significant main effect of time (t=47.75, p<.001), with higher average PDS at follow-up (mean=2.18, SD=0.68) compared to baseline (mean=1.67, SD=0.53), and a significant interaction between time and sex, indicating that females had significantly higher change in average PDS between the two assessments (β=0.33 vs β=0.17, p<.001).

### Brain age prediction accuracy

Figure 1C shows the distribution of brain age in females and males separately. Age was classified with a mean absolute error of 0.7 and 1.4 years at the two timepoints respectively, and an intraclass correlation of 0.65 between the two timepoints.

### The association between pubertal development and brain age

Table 1 and Figure 2 summarize the fixed effects results from models assessing main effects of pubertal development (hypotheses 1 and 2) on brain age. When controlling for the effect of age and sex (model 2), an increase of one stage in pubertal development was associated with a higher brain age of 2.4 months (β=0.10, p<.001), indicating a link between brain maturation and pubertal status across time. Further, a significant interaction effect between average PDS and timepoint (Model 3, p<.001) indicated a stronger association between puberty and brain age at follow-up than baseline.

**Table 1.**
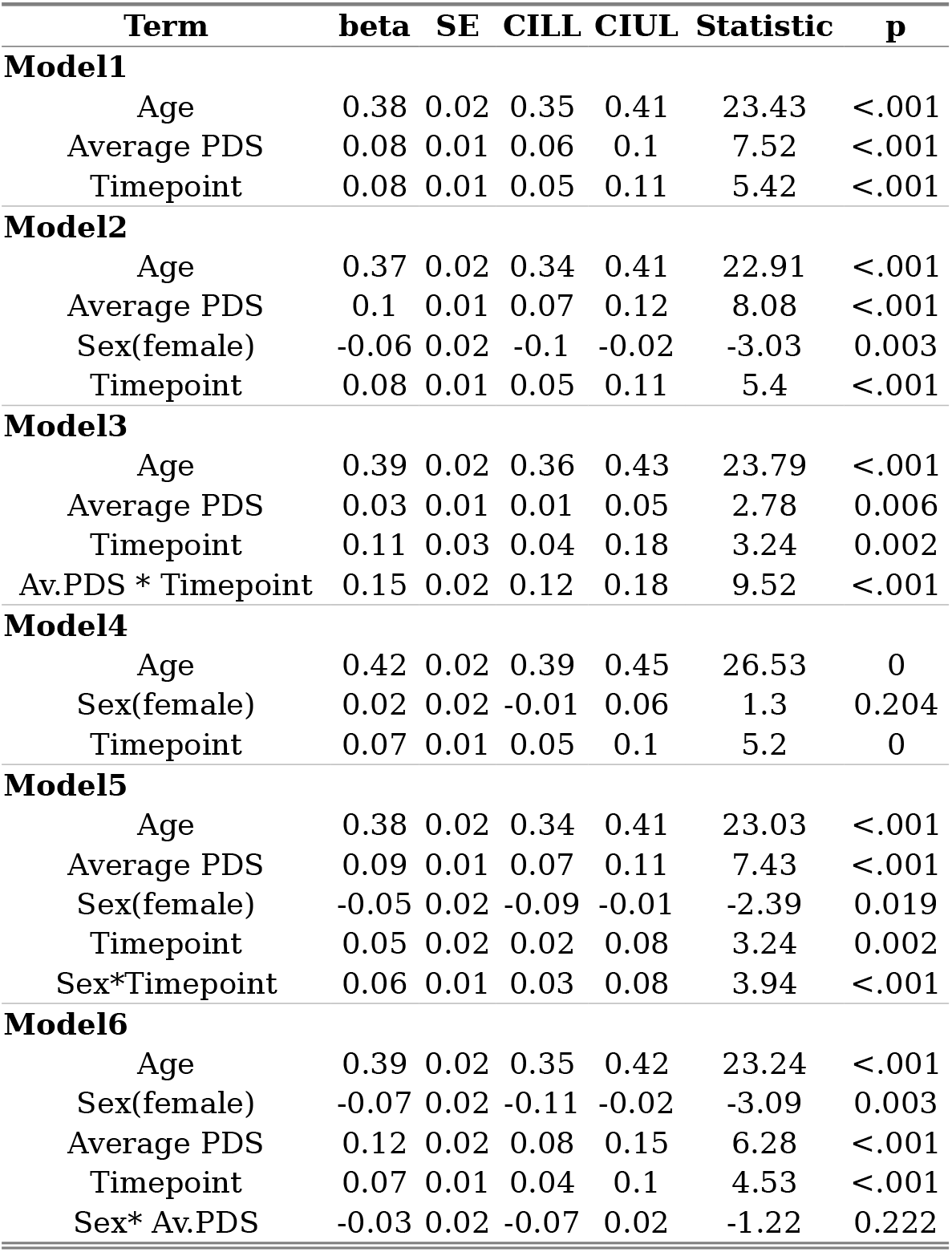
Results from linear mixed effects models on brain age LME output from all models including FDR corrected p-values and standardized beta coefficients.

**Figure 2.**
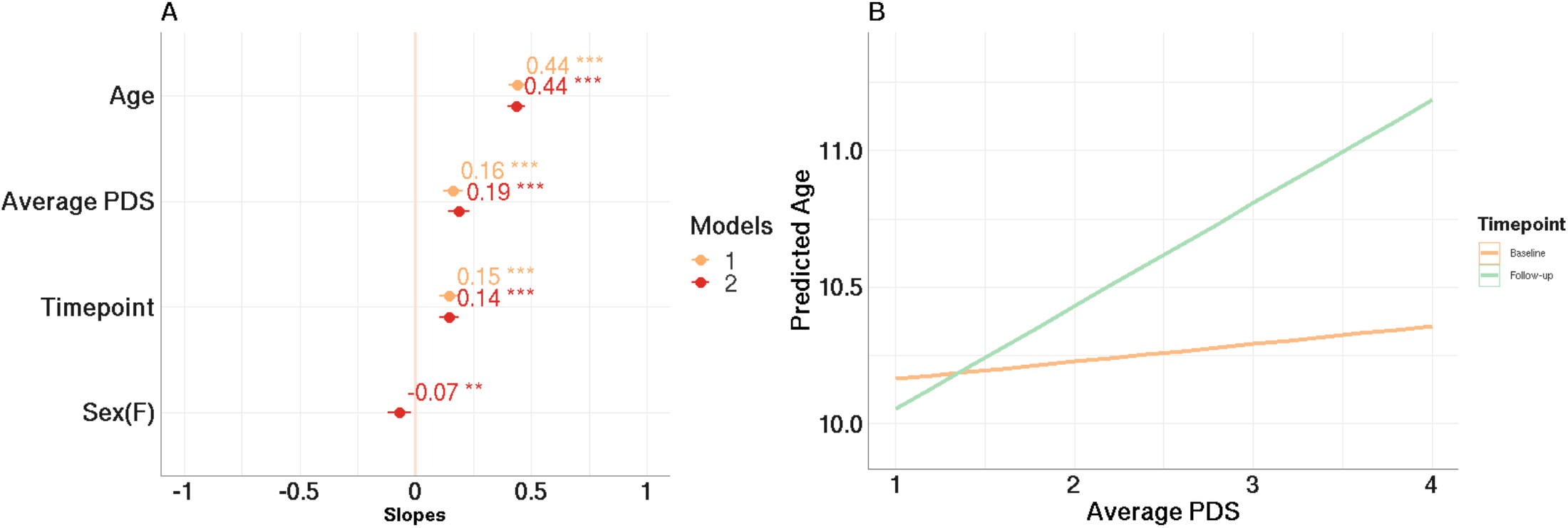
Associations between puberty and brain age. A: Standardized parameter estimates reflecting main effects obtained from model 1 and 2, with and without sex included in the model. The error bars reflect the 95% confidence interval. B: The interaction between average PDS and timepoint, indicating a moderating effect of puberty on brain age, in particular at follow-up.

### Sex differences in brain age

Table 1 and Figure 3 summarize the fixed effects from models assessing sex differences in brain age (model 3 and 4). When including average PDS in the models, females had a significantly lower brain age of almost one month across timepoints compared to males (Model 2, β=-0.06, p <.001). When omitting average PDS from the model, the sex differences in brain age were not significant (Model 4, β=0.02, p=.2).

**Figure 3.**
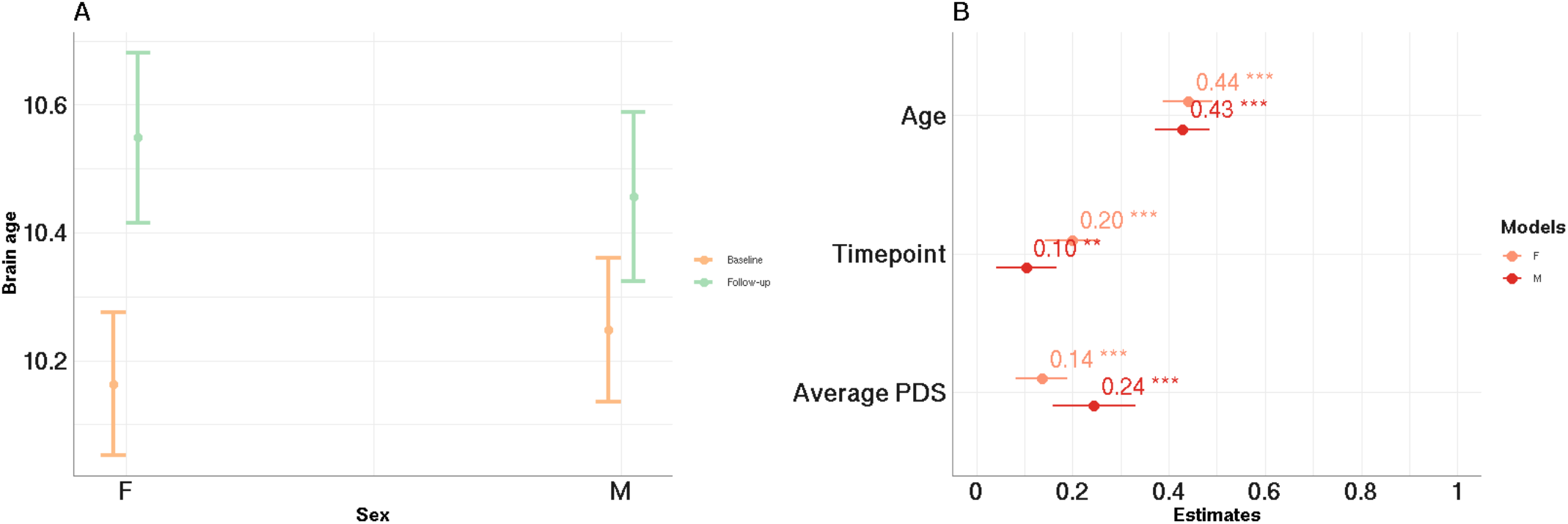
Sex specific effects of puberty and longitudinal development of brain age. A: Interaction between sex and time from model 5, showing larger longitudinal changes in brain age in females compared to males while controlling for average PDS. B: The relationship between average PDS, age, timepoint, and brain age within females and males, respectively (interaction not significant).

A significant interaction effect between sex and timepoint was observed when including average PDS in the model (model 5, p<.001), with follow up analysis indicating larger increase in brain age over time in females compared to males (β = 0.14, vs β = 0.08, Figure 3: C).

Sex-specific models revealed a significant relationship between pubertal development and brain age in females (β = 0.08, p<.001) and males (β=0.15, p< .001). The strengths of the associations were not significantly different, as indicated by a non-significant interaction term (Model 6, sex*average PDS, p=.22).

## Discussion

The age of pubertal onset has been linked to several real-life outcomes, including educational achievements and psychopathology. Puberty-related influence on brain maturation could represent a relevant explanatory or mediating factor, and could also provide a window to the study of sex differences in brain and behavior. In the current study longitudinal brain age prediction during a sensitive period of early adolescence revealed a positive association between parent-rated pubertal development and brain age, indicating a relationship between pubertal development and brain maturation over and beyond chronological age. Compared to males, females exhibited a more advanced and a faster pace of pubertal development and also a higher rate of changes in brain age between time points. When accounting for sex differences in pubertal development, females exhibited an overall slightly lower brain age than their male peers across timepoints, which was not evident when omitting pubertal status for the models.

### Pubertal development and brain age

The results from models assessing main effects of PDS on brain age showed that a one stage increase of averaged PDS score was related to a higher brain age of 2.4 months, when controlling for sex and age. This supports our first hypothesis of a contribution of pubertal development to overall brain maturation, in line with previous research linking pubertal development to morphological changes in brain structure (Vijayakumar et al., 2021; Goddings et al., 2014; Wierenga et al., 2018). Pubertal development also significantly interacted with time in relation to brain age, indicating that changes in brain age over time depends on pubertal stage and development. The effect sizes were moderate and comparable to previous studies using other neuroimaging-based outcomes to study the associations between brain structure and puberty (Vijakumar et al., 2021). Brain development during adolescence is likely shaped by an interaction of genetically programmed age-related changes, biological processes, and fluctuating environmental pressures (Fernandez-Cabello et al., 2022; Ferschmann et al., 2022). Combined with previous findings, our study shows that the effects of puberty are non-negligible and should be considered an influencing factor when studying adolescent brain development.

### Sex differences in adolescence brain age

The developmental processes shaping adolescent and adult sex differences in brain and behavior comprise a combination of hard-wired biological processes and complex sociocultural and environmental influences, and their dynamic interactions. The earlier pubertal onset and maturation among females has been proposed among the candidate mechanisms of sex differences in school performance and other life outcomes (Pfeifer & Allen, 2021; Torvik et al., 2021). While our analysis revealed a positive association between pubertal development and brain maturation across sexes, females showed higher rates of pubertal development and a higher rate of brain age increase between assessments. Models accounting for PDS revealed a main effect of sex on brain age, indicating an overall slightly lower brain age among female participants compared to their male peers across time points. This effect was driven by sex differences at baseline, since and female sex was associated with a slightly higher brain age at follow-up. Interactions between sex and time point indicated that males had a significantly lower increase in brain age between timepoints compared to females. This sex difference in the brain maturational tempo during puberty is notable and points towards an aspect of adolescent brain development of relevance for future research.

Our findings of overall lower brain age in female compared to male participants contrast the findings from a recent study including 330 youth aged 12-17 years reporting that female participants had a higher brain than their male peers (Brouwers et al., 2021). This discrepancy may be partly due to the different age ranges and level of maturation in the two samples. Indeed, the current findings revealed that females showed slightly higher brain age at follow-up, and their overall lower brain age was only observable when including pubertal development in the model. Thus, it is conceivable that the apparently divergent findings reflect sex differences in the rate of brain maturation occurring during puberty. Subsequent follow-up assessments of the ABCD cohort will be able to pursue this hypothesis, and may further characterize the involvement of pubertal development in the brain developmental processes laying the foundation of sex differences in complex traits and behaviors in adulthood.

Due to the surge in gonadal hormones, sex-differentiation of brain structure and function is believed to be largely shaped during puberty. Despite this, research findings are inconsistent regarding morphological brain differences emerging during this period (Lenroot & Giedd, 2010) and even in adulthood (Ritchie et al., 2021). Recent studies have reported higher inter-individual variability in brain structural volumes among males compared to females (Wierengia et al., 2018a; Wierengia et al., 2018b; Wierengia et al., 2019b), possibly indicating a wider range of influencing factors and larger individual differences in brain structure among male than female participants. The sex differences in brain age change seen in the current study may point to highly relevant sex differences in the onset and tempo of brain maturational processes and milestones. Moreover, the observed relationships between pubertal development and brain maturation in both sexes indicate that puberty is a key process to take into consideration when studying sex differences in adolescent neurodevelopment.

### Strengths and limitations

Key strengths of our study include the large number of subjects and the longitudinal aspect, enabling analysis of changes in brain age over a period of two years and sufficient power to reduce uncertainty of the estimates and detect relatively small effects.

The narrow age range in our sample may have provided more power to disentangle the effects of age and puberty on brain maturation. Age and pubertal development are highly correlated, and it has been suggested that studying a sample with a shorter age-range could disentangle the brain maturation attributable to age and pubertal development variability (Goddings et al., 2019). However, due to the young age of the sample we were not able to capture the complete developmental trajectory of brain age during the full course of puberty, as youths have just experienced the onset of puberty at this age. Thus, our results can be best described as capturing the effects of early pubertal development. Moreover, assuming that gonadal steroids is one of the biological factors driving the pubertal impact on brain development, there may be a time difference in their effect on neural properties compared to physiology, such that effect on brain properties are more detectable later in development. The young age and overall early pubertal status in our sample thus prohibits any analysis of potential temporal delay between the onset of puberty and its effect on brain maturation. Another challenge to the interpretation of the observed relationships between pubertal development and brain maturation is the possible confounding effects of socioeconomic factors such as poverty, air pollution and parental education and their complex interactions with genetic factors, both through direct and indirect effects (Bleile et al., 2017; Styne, 2004). Relatedly, future studies should test to which degree lifestyle and health-related behaviors such as physical activity, nutrition and obesity mediate the current association between puberty and adolescent brain development (Styne, 2004; Bleile et al., 2017; Beck et al., 2021).

Brain age was derived from a deep learning model without predefined regions of interests, providing an anatomically unbiased estimate of brain age. This might be advantageous in a young age sample as the neurodevelopmental trajectories during adolescence is heterogeneous and non-linear across individuals and brain regions (Østby et al., 2009). In this study the training and validation of the model was performed in a dataset with an age range spanning the full lifespan, while the analyses were performed in data spanning a narrow age-range during early adolescence. The differences in MAE accompanied by a difference in mean error of 0.6 years between the two timepoints indicates that the validation procedure might not have been sensitive enough to pick up strongly non-linear, local effects such as a sex specific age bias in the given age range. The general age bias was controlled for in LME models, and is not necessarily a problem.

Further, while the brain age model provided highly accurate estimates based on anatomically unbiased information, it was only informed by the signal embedded in the T1-weighted MRI data. It is possible that a multimodal approach integrating different imaging modalities could have provided brain age estimates with varying levels of sensitivity and specificity (Rokicki et al., 2021), which could offer the opportunity to triangulate different biological processes related to puberty and brain maturation. Further studies using a wider range of the rich neuroimaging data available in the ABCD study may be able to test this hypothesis.

A final limitation to our methods is the subjective nature of the pubertal assessment. Although parent rated PDS has shown high inter-rater reliability compared to clinician rated Tanner staging (Koopman-Verhoeff, 2020; Carskadon & Acebo, 1993), there might be variability in the parents/caregivers awareness of their children’s pubertal development that could not be controlled for. In sum, future studies will benefit from a wider age-span of subjects (and consequently more developed in puberty) when investigating the effects of puberty on brain maturation, as well as from objective assessment of puberty via blood plasma or saliva assessment of sex-steroids. Long-term follow-up assessments are required to assess the long-term real-life impact of individual differences in the onset and pace of puberty and its associations with brain maturation.

Taken together, this study suggests that pubertal development mediates overall brain maturation during adolescence. Although the sex differences in brain age were relatively small, females presented with more advanced and higher rates of changes in their pubertal development and also exhibited larger changes in brain age between baseline and follow up. Thus, our results indicate a link between the temporal characteristics of pubertal and brain development that may be provide a relevant window into the neurodevelopmental and neuroendocrinological origins of sex-differences in relation to mental health and other life outcomes.

## Data Availability

All data produced are available online at https://data-archive.nimh.nih.gov/abcd).

## Acknowledgements

The work was performed on the Service for Sensitive Data (TSD) platform, owned by the University of Oslo, operated and developed by the TSD service group at the University of Oslo IT-Department (USIT). Computations were also performed using resources provided by UNINETT Sigma2—the National Infrastructure for High Performance Computing and Data Storage in Norway. While working on this study, the authors received funding from the Research Council of Norway (273345, 249795, 298646, 300768, 223273), the South-Eastern Norway Regional Health Authority (2018076, 2019101), the European Research Council under the European Union’s Horizon 2020 research and innovation program (802998), and the Swiss National Science Foundation (PZ00P3_193658).

